# Threshold analyses on combinations of testing, population size, and vaccine coverage for COVID-19 control in a university setting

**DOI:** 10.1101/2020.07.21.20158303

**Authors:** Xinmeng Zhao, Hanisha Tatapudi, George Corey, Chaitra Gopalappa

## Abstract

We simulated epidemic projections of a potential COVID-19 outbreak in a residential university population in the United States under varying combinations of asymptomatic tests (5% to 33% per day), transmission rates (2.5% to 14%), and contact rates (1 to 25), to identify the contact rate threshold that, if exceeded, would lead to exponential growth in infections. Using this, we extracted contact rate thresholds among non-essential workers, population size thresholds in the absence of vaccines, and vaccine coverage thresholds. We further stream-lined our analyses to transmission rates of 5 to 8%, to correspond to the reported levels of face-mask-use/physical-distancing during the 2020 pandemic.

Our results suggest that, in the absence of vaccines, testing alone without reducing population size would not be sufficient to control an outbreak. If the population size is lowered to 34% (or 44%) of the actual population size to maintain contact rates at 4 (or 7) among non-essential workers, mass tests at 25% (or 33%) per day would help control an outbreak. With the availability of vaccines, the campus can be kept at full population provided at least 95% are vaccinated. If vaccines are partially available such that the coverage is lower than 95%, keeping at full population would require asymptomatic testing, either mass tests at 25% per day if vaccine coverage is at 63-79%, or mass tests at 33% per day if vaccine coverage is at 53-68%. If vaccine coverage is below 53%, to control an outbreak, in addition to mass tests at 33% per day, it would also require lowering the population size to 90%, 75%, and 60%, if vaccine coverage is at 38-53%, 23-38%, and below 23%, respectively.

Threshold estimates from this study, interpolated over the range of transmission rates, can collectively help inform campus level preparedness plans for adoption of face mask/physical-distancing, testing, remote instructions, and personnel scheduling, during non-availability or partial-availability of vaccines, in the event of SARS-Cov2-type disease outbreaks.

## Introduction

The COVID-19 pandemic caused by the SARS-CoV-2 virus has caused significant disease and economic burdens since its first outbreak in December 2019. Because of the absence of an effective vaccine, as of June 2020 at the time of this study and since March 2020, the main intervention for the prevention of COVID-19 transmissions had been to reduce contacts between people through lockdowns of non-essential organizations and services [1]. However, lockdowns are a threat to the economic stability of a nation as seen by the unprecedented rise in unemployment rates [2,3]. Therefore, while lockdowns are a good short-term strategy, for a long-term strategy, or until a vaccine becomes widely available, it has become necessary to identify alternate strategies and lifestyles that control the disease burden while minimizing the economic burden. Interventions that are effective include the use of face masks, physical distancing between persons at a recommended 6ft, and contact tracing and testing or mass testing to enable early diagnosis in the asymptomatic stage of infection [4]. However, removal of lockdowns should be strictly accompanied by a reopening plan that rapidly and efficiently enables the adoption of the above interventions to avoid an epidemic rebound. In addition to public health agencies, all members of a community, in both public and private sectors, play a key role in the development and implementation of a reopening plan that is most suited for their organization [5]. Among these sectors, universities and colleges bear a special burden to develop a reopening plan that include changes to a range of activities related to teaching, research, dining, housing, and extra-curricular activities.

We developed a compartmental differential equations model to simulate epidemic projections of a potential COVID-19 outbreak in a population of 38,000 individuals, which is representative of undergraduate and graduate students, faculty, and staff in a typical residential university in the United States. We simulated epidemic projections of potential outbreaks under varying combinations of contact tracing and testing, and mass testing, to identify combinations that would reduce the effective reproduction number *R*_*e*_ to a value below the epidemic threshold of 1. *R*_*e*_ is directly proportional to the duration of infectiousness, transmission rate (the probability of transmission per contact per day, representing the infectiousness of the virus), and contact rate (the number of contacts per person per day) [6]. Asymptomatic testing through trace and test or mass tests lead to diagnosis in the asymptomatic phase of the infection, and thus, if persons diagnosed with infection are successfully quarantined, it reduces the duration of exposure [7–9] and thus reduce *R*_*e*_. Physical distancing by the recommended 3 or 6ft and use of face masks can reduce transmission rate, and thus reduce *R*_*e*_ [10,11]. Reducing contact rate such as through transitioning to remote work to reduce population density on campus directly reduces *R*_*e*_. Thus, different types of interventions help reduce each of these components of *R*_*e*_. Here, we evaluated different combinations of test rates, transmission rates, and contact rates that help reduce *R*_*e*_ to below 1 to identify minimum levels of testing, physical distancing and face mask use, and population density necessary for effective control of an outbreak.

While it is generally known that increasing contact tracing and testing is necessary, studies evaluating testing at an organizational level, such as university, were only recently emerging at the time of this study in June 2020. One study that analyzed contact tracing in the general populations estimated that reducing *R*_0_ of 1.5 to an *R*_*e*_ of 1 requires more than 20% of contacts traced, reducing *R*_0_ of 2.5 to an *R*_*e*_ of 1 requires more than 80% of contacts traced, and reducing *R*_0_ of 3.5 to an *R*_*e*_ of 1 requires more than 100% of contacts traced [12]. A modeling study applied to the Boston area [13] estimated that the best way-out scenario is a Lift and Enhanced Testing (LET) with 50% detection and 40% of contacts traced. According to this, the number of individuals that need to be traced per 1000 persons is below 0.1 under partial reopening and below 0.15 under total reopening. Models for a university were only recently emerging at the time of this study in June 2020, [14–17] but typically, most studies combine transmission rate and contact rate as one metric in the evaluation of testing.

In this study, instead of using a product of transmission rate and contact rate as one metric as typically done, we evaluated these separately, due to the following reasons. First, it helps systematically evaluate different interventions considering that different types of interventions help reduce each of the three components of *R*_*e*_, testing reduces duration of exposed infectious stage, transitioning to remote classes reduce contact rate, vaccinations reduce the number of contacts who are potential disease carriers, and face mask use and 6ft distancing reduces transmission rate. Second, while adoption of each of these decisions are made at an organizational level, adherence and feasibility of face mask and 6ft distancing are highly influenced by individual behaviors and thus have a larger range of uncertainty. Third, while physical distancing and use of face masks can reduce transmission rate, the baseline transmission rate and expected reductions could vary based on multiple factors such as indoor vs. outdoor settings and ventilation, proper use and type of face mask [10,11,18,19], mode of transmission [20–23], and viral load in the index person [8,24]. Fourth, though we specifically focus this study on COVID-19 caused by the original SARS-CoV-2 virus, studying varying levels of transmission rates could help extrapolate findings to new variants or future outbreaks of viral respiratory infections with similar disease progressions [24], especially in the early stages when specific data is lacking but when the same non-pharmaceutical interventions, such as face masks, physical distancing, remote instructions, and testing, are suitable options.

To systematically inform these analyses, we first evaluated different combinations of trace and test rate, mass test rate, and transmission rate for a range of contact rates, to identify the threshold contact rates that maintain infection cases below certain set levels of tolerance. We then used the contact rate thresholds to identify the population size thresholds, i.e., the maximum population size on campus, which could help inform decisions related to campus activities such as the fraction of classes to transition to remote. We also used the contact rate thresholds to identify the vaccine coverage thresholds for a post-vaccine era, i.e., the vaccine coverage necessary for a campus to return to a normal population size. We also identify, under each intervention combination, the number of trace and tests and quarantines. These metrics could collectively help inform development of a preparedness plan for reopening a university during the COVID-19 pandemic or to set protocols in the event of future outbreaks.

## Methodology

### Simulation methodology

We developed a compartmental model for simulating epidemic projections over time. The epidemiological flow diagram for the compartmental model is depicted in Fig 1A. Each box is an epidemiological state, and each arrow represents a transition from one state to another. Note, each compartment is further split by age and gender, but for clarity of notations, we do not include it in the equations below.

Let π_*t*_ = [S, L, E, I, Q_L_, Q_E_, Q_I_, H, R, D] be a vector, with each element representing the number of people in a compartment at time *t*, specifically,

*S* = the number of susceptible individuals at time *t*,

*L* = the number of exposed, but asymptomatic and not infectious individuals (latent stage; also, the non-infectious phase of incubation stage) at time *t*,

*E* = the number of asymptomatic or pre-symptomatically infectious individuals (infectious phase of the incubation stage) at time *t*,

*I* = the number of infectious individuals (symptomatic and infectious stage) at time *t*,

*Q*_*L*_ = the number of exposed, asymptomatic and not infectious (latent) and quarantined individuals (diagnosed) at time *t*,

*Q*_*E*_ = the number of asymptomatic or pre-symptomatically infectious and quarantined individuals (diagnosed) at time *t*,

*Q*_*I*_ = the number of infectious and quarantined individuals (diagnosed) at time *t*,

*H* = the number of hospitalized individuals at time *t*,

*R* = the number of recovered individuals at time *t*, and

*D* = the number of deaths at time *t*.

**Fig 1.**
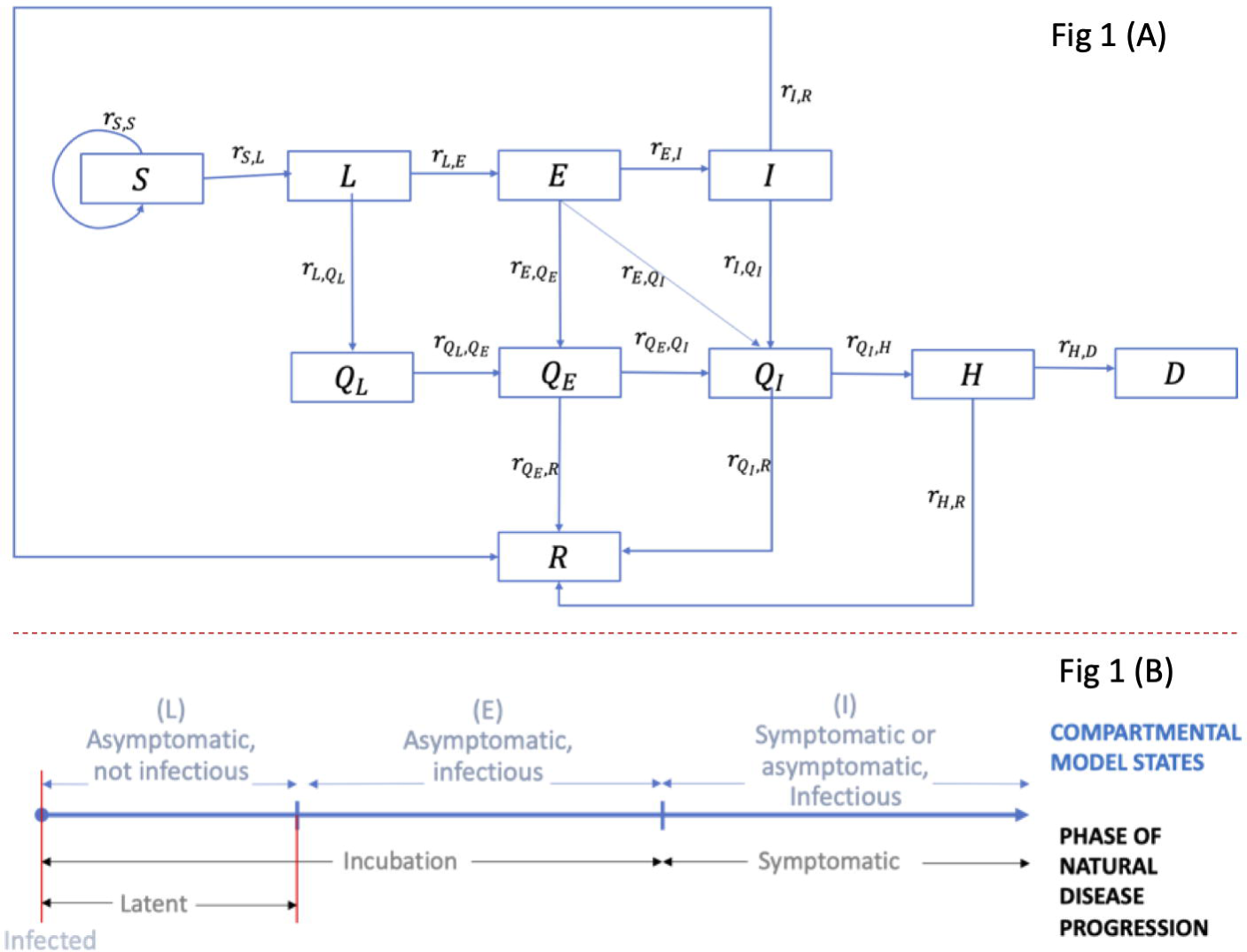
Overview of the extended SEIR compartmental model. (A) Compartmental model flow diagram. (B) Natural disease progression of SARS-COV-2 virus in infected patients. S = susceptible, L = exposed and not infectious (Latent stage) (asymptomatic), E = asymptomatic and infectious, I = symptomatic and infectious, *Q*_*L*_ = exposed and not infectious (Latent) and Quarantined (diagnosed), *Q*_*E*_ = asymptomatic and infectious and Quarantined (diagnosed), *Q*_*I*_ = Infectious and Quarantined (diagnosed), H = Hospitalized, R = Recovered, and D = Deaths.

Epidemic states *L, E*, and *I* were formulated such that each state represented a distinct phase along the natural disease progression (see Fig 1B), and they collectively included all phases. Over time, persons from *S* can transition to *L, E*, and *I*, and upon diagnoses, transition to *Q*_*L*_, *Q*_*E*_, or *Q*_*I*_, and further to *H, R*, or *D*, (transitions represented by arrows in Fig 1A) as discussed below.

Let,

*p* = transmission rate (probability of transmission per contact per day),

*c* = contact rate (number of contacts per person per day),

*N* = total population who are alive,

*a*_*B*_ = symptom-based testing rate,

*a*_*C,t*_ = rate of testing through contact tracing at time t,

*a*_*U,t*_ = rate of testing through mass testing at time t,

*ρ* = test sensitivity for asymptomatic testing (through mass tests or trace and test),

*days*_*L*_ = duration in latent period,

*days*_*incub*_ = duration in incubation period,

*days*_*IR*_ = time from onset of symptoms to recovery,

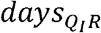 = time from diagnosis to recovery,

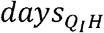 = time from diagnosis to hospitalization,

*days*_*HR*_ = time from hospitalization to recovery,

*days*_*HD*_ = time from hospitalization to death,

*prop*_*hosp*_ = proportion hospitalized, and

*prop*_*severe*_ = proportion of cases that are severe.

Then, we can write the equations for transition rates (arrows in Fig 1A) as follows:

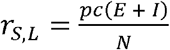, which assumes that only infected persons in *E* and *I* can transmit, persons in *Q*_*E*_ and *Q*_*I*_ self-quarantine, and persons in *L* and *Q*_*L*_ are not infectious.

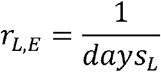

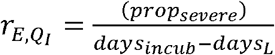, which assumes that only a proportion of cases that are severe (*prop*_*severe*_) get diagnosed immediately because of exhibition of symptoms, we use the proportion hospitalized as a proxy for severe cases; the denominator is based on the assumption that the duration in state E is equal to the difference between the duration of the incubation period and the latent period.

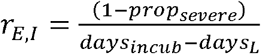, which follows from above.

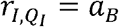, which assumes that under symptom-based testing, only persons who show moderate to severe symptoms get diagnosed and those who show mild symptoms do not.

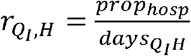, for *prop*_*hosp*_ we use the proportion of persons hospitalized among those diagnosed through symptom-based testing.

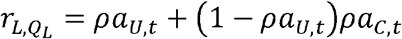, which assumes that under the implementation of both mass testing and contact tracing and testing, persons diagnosed through mass test will not be tested again on the same day through contact tracing (as our time unit is daily).

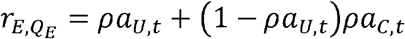, which is similar to above.

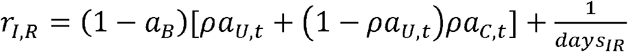, which assumes that persons with mild cases that did not get diagnosed through symptom-based testing have a chance of getting tested through additional testing options, and self-quarantine upon diagnosis. Note that we did not separately model asymptomatic cases but incorporated that into the symptom-based testing rate (*a*_*B*_) by considering that 35% of cases are mild to no symptoms and thus do not have a chance of being diagnosed through symptom-based testing.

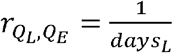

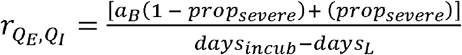, theoretically, 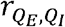 should be the same as *r*_*E,I*,_ however, as the rate of transitioning from *Q*_*I*_ to *H* is fixed to fit to the proportion hospitalized under symptom-based tests, if extensive testing is conducted, the number of persons in *Q*_*I*_ would increase, thus, incorrectly inflating the number of persons who are hospitalized; To avoid this, we modified the equation to consider that the number of persons flowing into *Q*_*I*_would be equal to the proportion flowing from *I* to *Q*_*I*_ under symptom-based testing.

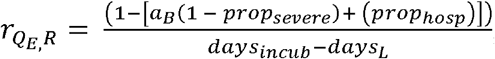, which follows from the above equation.

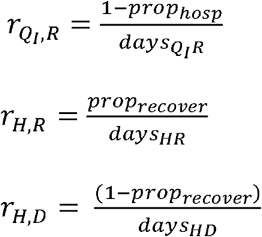

Note: r_s,s_ is the testing rate (either through mass test or trace and test). We assumed that susceptible persons go back to the susceptible state after testing, i.e., we did not explicitly track false positives.

The values and ranges for the above epidemic parameters used in the compartmental simulation model are presented in Supplemental Appendix Table S1.

We simulate the epidemic over time using the following system of differential equations

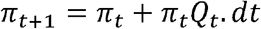

where, *Q*_*t*_ = a matrix of transition rates between states (arrows in Fig 1A), and *dt* = time-step. We use a time-unit of per day for the transition rates in *Q*_*t*_ and set 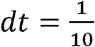, and thus, the model simulates every 10^th^ of a day.

The expansion of the system of differential equations are as follows:

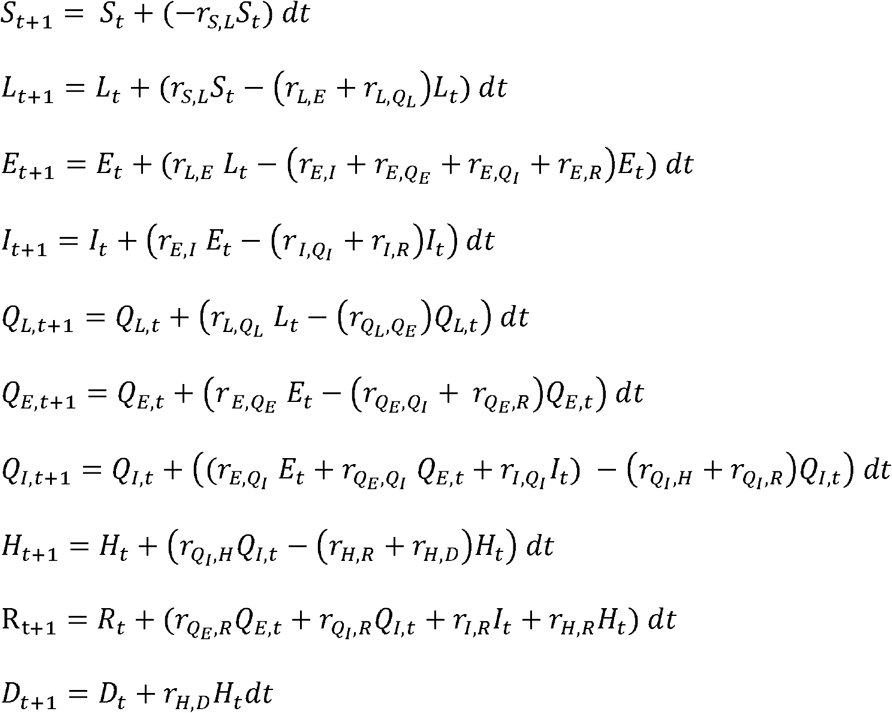

We can further expand by substitution of the rate terms with their equations as follows:

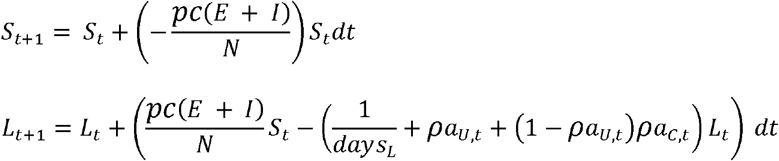

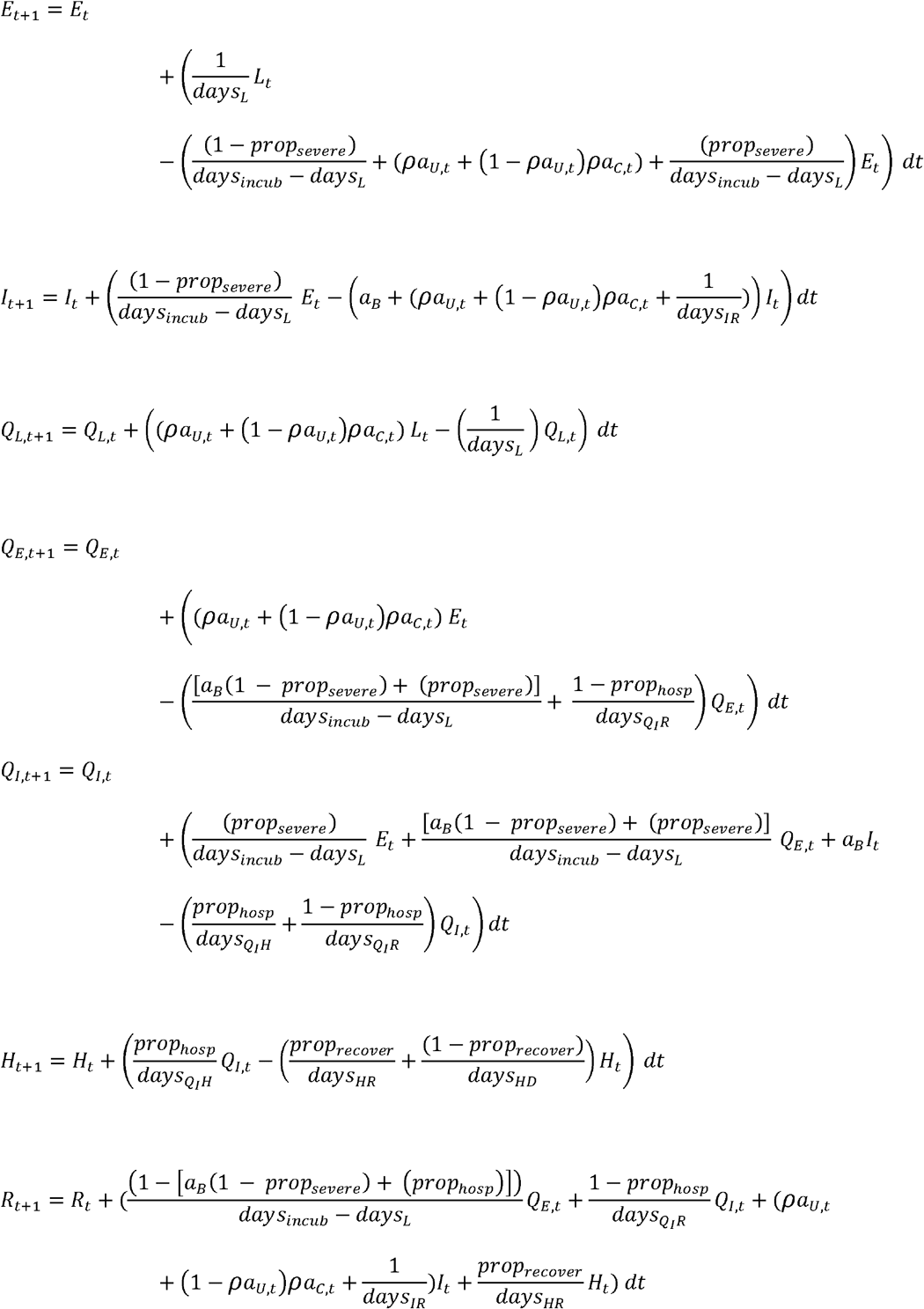

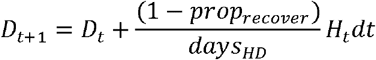

### Input data assumptions and sources for simulation model

For the rates of natural disease progression, we used estimates from other studies in the literature. The description of the data, sources, and values (with ranges and medians where applicable) for all parameters are available in the Supplemental Appendix Table S1. Briefly, we assumed an incubation period duration of 5.4 days [25], the first 2.5 days in stage L (not infectious and asymptomatic) [26], and the remaining 2.9 days in stage E (infectious and asymptomatic). We assumed about 65% of cases develop medium to severe symptoms [27] and, in the absence of test and trace or mass test, can be diagnosed through symptom-based testing. We assumed the remaining 35% of cases show mild to no symptoms and can be diagnosed only through trace and test, or universal mass test. We assumed an average duration of 3.5 days from the time of onset of symptoms to hospitalization [28], with the proportion hospitalized varying as a function of age. For mild cases, we assumed an average duration of 7 days from the time of onset of symptoms to recovery [28]. We assumed case fatality rates vary as a function of age and gender.

### Interventions

#### Mass test and trace and test

We evaluated the following scenarios: mass test only, trace and test only, delayed trace and test only, combination mass test and trace and test, and combination mass test and delayed trace and test, each at different rates of testing, as follows. We evaluated mass testing at rates of 5% 10%, 20%, 25%, and 33% of the population per day, which is equivalent to testing once every 20 days (5% per day over every 20-day period), 10 days, 5 days, 4 days, and 3 days (33% per day every 3-day period), respectively. We modeled the rate of trace and test as the inverse of the time from infection to effective isolation of a contact, i.e., the sum of the number of days passed since contact with an individual (as reported by the index diagnosed person) and the number of days into the future to find, test, and isolate the infected contact. We chose this definition as each component in this duration can vary significantly for every diagnosed person and for each of their contacts. In the case the contact is never found, the duration would be the full duration of infection. Thus, this definition of trace and test can be comparable to data that is typically collected. Specifically, the trace and test rate here should be compared to the average of the inverse of the time from reported contact to either effective isolation of that contact or maximum infection duration (whichever is the least value), averaged over all contacts. We evaluated trace and test rates at levels of 10%, 17%, 20%, 25%, 33%, and 50%, equivalent of 10 days, 6 days, 5 days, 4 days, 3 days, and 2 days, respectively, from the time of transmission of infection to effective isolation of that contact. We evaluated combinations of mass test and trace and test, by varying mass tests between 5% and 33% per day and keeping trace and test at 50% as this higher rate of trace and test maybe more feasible with mass test than symptom-based test only. We assumed trace and test would initiate within the first 5 cases of diagnosis. Considering there may be delays in setting up a trace and test system (such as in events of new outbreaks in the future or failure to respond quickly), to tests its sensitivity, we evaluated scenarios by delaying the initiating of trace and test to after diagnosis of 20 cases. Thus, the scenarios referred to as ‘trace and test only’ and ‘combination tests’ refers to initiation of trace and test after first 5 cases of diagnosis. And the scenarios referred to as ‘delayed trace and test only’ and ‘delayed combination tests’ refers to delaying initiation of trace and test to after diagnoses of 20 cases. In all scenarios, we applied baseline symptom-based testing, assumed test results are available within 24 hours, and persons testing positive self-quarantine for 14-days. For diagnosis in asymptomatic stages, i.e., diagnosis through trace and test or mass test, we assumed a test sensitivity of 0.9 [29].

#### Non-pharmaceutical interventions

We evaluated transmission rates (*p*) of 14% (baseline), 8% (mid), 5.4% (lower-mid), and 2.5% (lowest). The baseline value of *p* corresponds to an average estimate under no interventions (no physical distancing and no face masks) [11,30]. A transmission rate of 8% corresponds to the expected rates with the use of face masks in a non-health care setting [11]. Transmission rates of 5.4% and 2.5% correspond to expected rates under 3ft and 6ft physical distancing, respectively [11] (see Supplemental Appendix Table S2). We evaluated contact rates between 1 and 25 (*c*), we did not differentiate between on-campus and off-campus contact rates.

### Application to a university setting

#### Demographic data

We used the Fall 2018 student enrollment data from the University of Massachusetts - Amherst, Amherst, MA, to determine the population size of undergraduate and graduate students and their age and gender distributions [31]. For faculty and staff, we used the age distribution of persons 25 years and older from the Town of Amherst, MA, where the university is located [32]. To initiate an outbreak, we assumed 4 to 5 infected cases on Day 1, estimated as follows. We assumed that the proportion of incoming students who are infected would be equal to the prevalence of COVID-19 in Massachusetts (MA) in June 2020. We also assumed that all incoming students would be tested, and about 10% of infected cases would be false negatives. Prevalence is unknown, as not all cases are diagnosed and diagnosed cases are not specifically tracked. Therefore, to estimate prevalence of COVID-19 in MA, we used the simulation model to determine the ratio of new diagnosis to persons with infection and applied that ratio to the number of new diagnoses on June 26^th^ in MA. This resulted in about 4 infected cases on day 1 remaining undetected, thus initiating an outbreak. We also assumed that at the beginning of every week, there would be about 3 to 4 infections from outside, calculated by assuming that about 10% of the population are likely to mix with the population outside the university or travel out of Amherst during weekends and are not tested upon return. Based on the above, we initialized the model on Day 1 with 4 infected persons in the Latent stage and added 3 to 4 outside cases to the Latent stage at the beginning of every week. We simulated the model for a 90-day period to represent the duration of the expected Fall 2020 semester.

### Tolerance on the number of infected cases for identifying contact rate thresholds

We evaluated contact rate thresholds under three levels of epidemic tolerance: relaxed tolerance, medium tolerance, and tight tolerance. Relaxed tolerance marked the point beyond which there was an exponential growth in infections, the maximum number of infections under this tolerance level was about 170. For medium tolerance, we set the number of infections to less than 77, and for tight tolerance, we set the number of infections to less than 50. The latter two cases correspond to maximum infections for a case fatality rate (CFR) of 2%, which is the reported CFR in the general population for the United States [33]. That is, 1/0.02 gives the 50 cases threshold and 77 is obtained by further dividing that by 65%, which is the proportion of cases with medium to severe symptoms [27], to account for the remaining 35% of cases with mild to no symptoms that were likely unreported and thus not included in the CFR calculation. As the CFR for COVID-19 is much lower in university student aged populations, the use of the alternative tolerance on the number of infections helps avoid spill-over effect of a breakout into the community. Also note that, because of our assumptions for the number of initial cases and cases per week entering the population from outside, the minimum number of cases over the 90-day period would be 45. Therefore, the tolerance of 50 cases is a very tight tolerance. For context, one of the indicators CDC uses to categorize community transmission risk is the number of cases per 100,000 persons during the last 7 days, categorizing as low, moderate, and substantial to high if there were less than 10, 10-49, and greater than 49 cases, respectively [34]. Converting our tolerance levels to the CDC indicator would translate to 35, 15, and 10 cases for relaxed, medium, and tight tolerances, respectively. If we exclude the 45 cases from outside, it translates to 25, 6, and 1 cases for relaxed, medium, and tight tolerances, respectively.

### Population behavioral data

While there was limited data on contact rates specific to university students at the time of this original study in June 2020, studies conducted since then have generated some (though limited) data on population behaviors. These data include contact rates and behaviors related to use of non-pharmaceutical interventions such as face mask and 6 ft physical distancing, mostly either self-reported in surveys or estimations made in other modeling studies informed through university settings. We briefly summarize the data from each study in the Supplemental Appendix Table S3. Some of the surveys were specific to university students in the United States while others were either university students in other countries or general populations. Studies on surveys of university students, when partial shutdowns were enforced and universities resorted to varying levels of remote classes, reported 6 to 8 contacts per person per day [35,36]. However, students who self-reported as providing essential services or caring for non-household members (∼23%) reported an average contact rate of about 14 per person per day [37]. Modeling studies that estimated contacts among university students for a scenario prior to the pandemic assume contact rates of 16 to 24 per person per day [38–40]. Using data on face mask use and physical distancing, specifically originating from three surveys of student and general population in the United States [38,42,43] and the transmission rates corresponding to these interventions (summarized in Supplemental Appendix Table S3), we calculate the expected transmission rate to be between 5% and 8%. We use these estimates to further streamline our analyses.

### Interpretation of contact rate thresholds: size of social circle, population size, and vaccine coverage

We utilize the contact rate thresholds, under the different levels of testing and transmission rate (face mask use and physical distancing), to identify four additional metrics that would help inform campus decisions: first, the contact rate threshold among non-essential workers after accounting for the higher contact rate among essential workers, which would help inform the size of social circles at the individual level and schedule campus activities; second, the threshold values for population size on campus as a proportion of the actual population size, which would help decisions related to the fraction of remote vs. face-to-face classes, on-campus housing, and other campus activities for the era of pre-vaccine availability; third, for the era of post-vaccine availability, the threshold values for vaccination coverage for the university to return to normal (i.e., 100% population size); fourth, the threshold values for population sizes under varying levels of vaccine coverage, which would help decisions related to campus activities in the event that vaccines are only partially available that coverage is not at levels sufficient to fully return to normal. All four metrics would be used alongside decisions related to the level of testing.

The metrics were estimated as follows. Suppose *Ĉ* is the contact rate threshold, we estimated the first metric as *Ĉ*_*n*_ = (*Ĉ* − *C*_*e*_*p*_*e*_)/(1 − *p*_*e*_), where *Ĉ*_*n*_ is the contact rate threshold among non-essential workers (we limit 0 ≤ *Ĉ*_*n*_ ≤ *Ĉ*), *C*_*e*_ is the contact rate among essential workers (we assume *C*_*e*_ =14 [37]), and *p*_*e*_ is the proportion of the population who are essential workers (we assume *p*_*e*_ =23% [37]).

The interpretation of the second, third, and fourth metrics arise from our simplifying assumption that contact rates are directly proportional to the population density [41], *C*=*c*_0_*ρ*; *ρ* = *N*/*A*; where *C* is the actual contact rate (under regular face-to-face instructions), *ρ* is the density, *N* is the population size, *A* is the campus area, and *c*_0_ is a constant. Further, we assume that university campuses maintain similar levels of population density under regular work conditions, i.e., though the population sizes may vary across universities, the campus area also changes proportionally so that the population density is similar, and thus, the contact rates under regular work conditions are also similar. Multiple studies reported similar contact rates of 16 to 24 under regular working conditions supporting this assumption [42,43]. Thus, if our estimated contact rate thresholds (say *Ĉ*) are lower than the actual contact rates of 16 to 24 (*C*), given fixed area (*A*), achieving *Ĉ* would require reducing the population size (*N*) proportional to the reduction in contact rate, i.e., 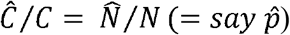, implying that the population size on campus should be at a maximum of 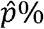 of its original population size 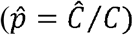.

The third metric on interpretation of threshold for vaccination coverage (say 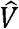) follows from the above assumptions. Achieving a contact rate threshold of *Ĉ* when universities are back to regular face-to-face classes, i.e., 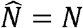 or 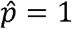, would require that (1 − *Ĉ* / *C*) proportion of the population be effectively vaccinated. More precisely, vaccine coverage should be at least 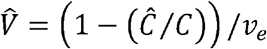, where *v*_*e*_ is vaccine efficacy and corresponds to the chance that a vaccinated individual is fully protected from being infected, and thus, is not a potential disease carrying contact. Intuitively, this is saying that though the actual contact rate is *C*, because 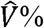 are vaccinated and protected from infection or transmitting, the effective contact rate is *Ĉ*. This implies that a threshold contact rate of *Ĉ* can be achieved, while maintaining 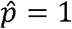, if 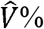 are vaccinated. The vaccine coverage results presented here were estimated by assuming a vaccine efficacy of 95%, and thus, in the event that this changes, the vaccine coverage results should be adjusted by multiplying with 95% and dividing by the new value.

Following from above, the fourth metric considers the fact that if the actual vaccine coverage (say *V*) is less than 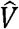, achieving the contact rate threshold (*Ĉ*) would also require some reduction in *N*. Specifically, the population size on campus should be at a maximum of 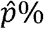 of its original population size, with 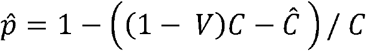, derived as follows. We can write (1 − *V*) *C* − *Ĉ* as the number of excess contacts, i.e., the number to reduce after accounting for the proportion vaccinated ((1− *V*) *C*), the proportion of contacts to reduce would then be ((1− *V*) *C* − *Ĉ*)/ *C*, and finally, applying the same assumptions as in the second metric would give the equation for 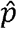. If the vaccination coverage is zero, i.e., *V* = 0, we would get back 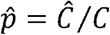. If *V* = 1, then 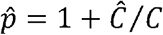, which implies that even if *Ĉ* = 0, the campus can fully open. We bound 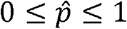, such that, even if *Ĉ* > 0 we interpret this as fully back to normal population size (though it would mathematically imply that the campus can handle a higher density from an epidemic perspective, e.g., influx from outside).

Thus, to keep within the infection tolerance levels, *Ĉ* would mark the maximum average contact rate over the full population, *Ĉ*_*n*_ the maximum average contact rate for non-essential workers after accounting for the higher contact rate among essential workers, 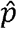 the maximum proportion of the population who should return back to campus (either when *V* = 0 or 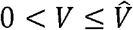), and 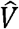 the minimum vaccine coverage to fully return back to normal 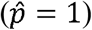. As the above method of estimation of thresholds incorporate the effectiveness of vaccinations, we can interpret that the interventions, such a testing and use of facemask and social distancing, would be applied to only the unvaccinated persons.

### Identifying feasible intervention combinations

We identify three sets of feasible combination results. For the event that vaccines are unavailable, we identify the feasible combinations of testing, contact rate for non-essential workers (*Ĉ*_*n*_), and population size on campus 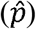 that can effectively control an outbreak to below the tolerance levels. We define feasible combinations as those with *Ĉ*_*n*_ > 2 in the transmission rate range of 5% to 8%, which would correspond to the reported use of face mask and physical distancing among the university population (see ‘Population behavioral data’ above). For the event that vaccines are partially or fully available, we identify the minimum vaccine coverage threshold 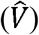 for the campus to fully return back to normal 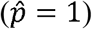, and if the vaccine coverage is below this threshold, the reductions in population size 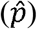 necessary to control the epidemic to within the tolerance levels. We also identify suitable testing scenarios for reported levels of face mask use and physical distancing (transmission rate of 5% to 8%), and reported levels of contact rates under regular face-to-face classes (16 to 24 per day) and remote classes (6 to 8 per day). We define suitable as those that avoid exponential growth in cases over the duration of a semester. For the above three sets of combination scenarios, we also present results under the full range of transmission rates in the Appendix Tables S4, S5, and S6, which could be useful in the event of change in transmission rates such as emergence of new virus variants.

## Results

When vaccines are unavailable (*V* = 0%), there is no single intervention that can effectively control an outbreak. However, there are multiple feasible combinations of testing, contact rate for non-essential workers (*Ĉ*_*n*_), and population size on campus 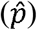 that can be implemented to effectively control an outbreak to keep cases below the relaxed to medium tolerance levels, though none to keep cases below the tight tolerance level (Table 1). Examples of feasible combinations under the relaxed tolerance level include: mass tests only at 25% per day, contact rate for non-essential workers at 2 to 6 per day, and campus population size at 26% to 42%; or trace and test only at 33%, contact rate for non-essential workers at 4 to 8 per day, and campus population size at 31% to 47% (see full list in Table 1). Under the medium tolerance level, only scenarios with combination tests were feasible, examples include: 5% mass test, 50% trace and test, contact rate for non-essential workers at 2 to 5 per day, and campus population size at 26% to 36%; or 33% mass test, 50% trace and test, contact rate for non-essential workers at 8 to 14 per day, and campus population size at 47% to 73% (see full list in Table 1). Note: the range in population size results correspond to mid-points of the range for contact rate () of 16 and 24 in Table 1.

**Table 1.** Feasible combinations ^□^ of testing, contact rate, and population size on campus for effective control of a disease outbreak in the absence of a vaccine.

| Tolerance | Testing | Contact rate threshold (per day)<sup>†</sup> for non-essential workers<sup>‡</sup> | Population size<sup>†</sup> (if regular contact rate is 16) | Population size<sup>†</sup> (if regular contact rate is 24) | Peak trace and tests per day (per 10,000 persons) | Peak quarantine per day (per 10,000 persons) |
| --- | --- | --- | --- | --- | --- | --- |
| Relaxed tolerance | S+25%U | 2 - 6 | 31% - 50% | 21% - 33% | 0 - 0 | 6 - 7 |
|  | S+33%U | 5 - 9 | 44% - 63% | 29% - 42% | 0 - 0 | 7 - 7 |
|  | S+33%T | 4 - 8 | 38% - 56% | 25% - 38% | 14 - 21 | 18 - 18 |
|  | S+50%T | 6 - 11 | 50% - 75% | 33% - 50% | 36 - 55 | 22 - 23 |
|  | S+5%U+50%T | 10 - 16 | 69% - 100% | 46% - 67% | 33 - 48 | 20 - 20 |
|  | S+10%U+50%T | 11 - 18 | 75% - 100% | 50% - 75% | 24 - 35 | 15 - 15 |
|  | S+20%U+50%T | 15 - 22 | 94% - 100% | 63% - 92% | 16 - 25 | 13 - 13 |
|  | S+25%U+50%T | 16 - 24 | 100% - 100% | 67% - 100% | 14 - 20 | 13 - 12 |
|  | S+33%U+50%T | 18 - 25 | 100% - 100% | 75% - 100% | 12 - 15 | 10 - 9 |
|  | S+5%U+50%dT | 4 - 9 | 38% - 63% | 25% - 42% | 36 - 55 | 21 - 25 |
|  | S+10%U+50%dT | 6 - 13 | 50% - 81% | 33% - 54% | 32 - 63 | 21 - 25 |
|  | S+20%U+50%dT | 11 - 18 | 75% - 100% | 50% - 75% | 36 - 54 | 21 - 21 |
|  | S+25%U+50%dT | 13 - 19 | 81% - 100% | 54% - 79% | 28 - 41 | 19 - 18 |
|  | S+33%U+50%dT | 15 - 23 | 94% - 100% | 63% - 96% | 21 - 33 | 16 - 18 |
| Medium tolerance | S+5%U+50%T | 2 - 5 | 31% - 44% | 21% - 29% | 8 - 9 | 6 - 6 |
|  | S+10%U+50%T | 4 - 8 | 38% - 56% | 25% - 38% | 7 - 11 | 6 - 6 |
|  | S+20%U+50%T | 5 - 10 | 44% - 69% | 29% - 46% | 4 - 7 | 5 - 5 |
|  | S+25%U+50%T | 6 - 11 | 50% - 75% | 33% - 50% | 4 - 6 | 4 - 5 |
|  | S+33%U+50%T | 8 - 14 | 56% - 88% | 38% - 58% | 3 - 6 | 5 - 5 |
|  | S+20%U+50%dT | 2 - 5 | 31% - 44% | 21% - 29% | 4 - 4 | 6 - 6 |
|  | S+25%U+50%dT | 4 - 6 | 38% - 50% | 25% - 33% | 4 - 4 | 6 - 5 |
|  | S+33%U+50%dT | 5 - 9 | 44% - 63% | 29% - 42% | 3 - 4 | 6 - 5 |
| Tight tolerance | No scenarios were feasible ☐ |  |  |  |  |  |

Relaxed tolerance: Less than 1 death or 170 cases of infection. This point also marks the point beyond which there was an exponential growth in infections in the simulated runs.
Medium tolerance: Less than 77 cases of infections. Estimated as $1/\text{CFR} \times \text{reported cases}$ . We assumed a case fatality rate (CFR) of 2% in the general population in the US [33]; We assumed that 65% of infected cases are reported, which is the proportion showing medium to severe symptoms [27].
Tight tolerance: Less than 50 cases of infection. Estimated as $1/\text{CFR}$ . We assumed a case fatality rate (CFR) of 2% in the general population in the US [33].
S: symptomatic testing, U: Mass test, T: trace and test, dT: delayed trace and test.
☐ We defined a testing scenario as feasible if estimated contact rate thresholds among non-essential workers were at least 2 when transmission rates were 8% and 5% (corresponding to reported use of face mask and physical distancing [11,30]). The range of values in the table thus correspond to transmission rate of 8% - 5%
<sup>‡</sup> We assume 23% are essential workers and have a contact rate of 14 per day [37].
<sup>†</sup> Contact rate threshold (per person per day): the average value for contacts per person per day to keep infections below the tolerance level. These reduced contact rates, from the original rates of 16 to 24 [42,43], can be achieved through reduction in population size at the noted thresholds.

The corresponding peak numbers of trace and tests per day (per 10,000 persons) in the above feasible scenarios were at a reasonably manageable level. The relaxed tolerance level had a higher value (14 to 55 per day) than the medium tolerance level (3 to 11 per day) considering the population size on campus were lower in the latter case because of the tighter tolerance (Table 1). The peak number of quarantines per day (per 10,000 persons) for the above feasible scenarios also seem manageable. As with above, the relaxed tolerance level had a higher value (6 to 25 per day) than the medium tolerance level (5 to 6 per day). Combinations of testing, contact rate, and population size for the full range of transmission rates evaluated are presented in Supplemental Appendix Table S4.

When vaccines become partially or fully available, to keep the population size on campus at 100% 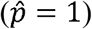, the level of testing necessary to effectively control an outbreak would depend on the vaccine coverage in the population (Table 2). To keep infection cases within the relaxed tolerance level, implementing symptomatic-testing-only will be sufficient if at least 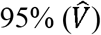 of the population are vaccinated (Table 2). With the addition of mass tests only, 5%, 10%, 20%, 25%, and 33% mass tests per day would be sufficient if at least 89% to 95%, 84% to 89%, 74% to 84%, 63% to 79%, and 53% to 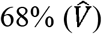 of the population are vaccinated (Table 2), respectively, the range corresponding to transmission rate of 5% to 8%, i.e., the unvaccinated continue to use face masks and maintain physical distancing at current compliance levels.

**Table 2.** Combinations of testing, vaccine coverage, and population size for effective control of a disease outbreak.

| Testing | Vaccination coverage | Population size (if regular contact rate is 16) | Population size (if regular contact rate is 24) |
| --- | --- | --- | --- |
| S | 95% - 100% | 100% - 100% | 100% - 100% |
| Mass tests only (% tested per day) |  |  |  |
| S+5%U | 89% - 95% | 100% - 100% | 100% - 100% |
| S+10% U | 84% - 89% | 100% - 100% | 100% - 100% |
| S+20%U | 74% - 84% | 100% - 100% | 100% - 100% |
| S+25%U | 63% - 79% | 100% - 100% | 100% - 100% |
| S+33%U | 53% - 68% | 100% - 100% | 100% - 100% |
| S+33%U | 38% - 53% | 100% - 97% | 79% - 83% |
| S+33%U | 23% - 38% | 85% - 82% | 64% - 68% |
| S+33%U | 8% - 23% | 70% - 67% | 49% - 53% |
| Trace and tests only |  |  |  |
| S+10%T | 84% - 95% | 100% - 100% | 100% - 100% |
| S+17%T | 79% - 89% | 100% - 100% | 100% - 100% |
| S+20%T | 74% - 84% | 100% - 100% | 100% - 100% |
| S+25%T | 68% - 84% | 100% - 100% | 100% - 100% |
| S+33%T | 58% - 74% | 100% - 100% | 100% - 100% |
| S+50%T | 42% - 63% | 100% - 100% | 100% - 100% |
| S+50%T | 27% - 42% | 100% - 92% | 77% - 75% |
| S+50%T | 12% - 27% | 87% - 77% | 62% - 60% |
| Trace and tests only (capped at 20%) |  |  |  |
| S+10%T | 84% - 95% | 100% - 100% | 100% - 100% |
| S+17%T | 79% - 89% | 100% - 100% | 100% - 100% |
| S+20%T | 74% - 84% | 100% - 100% | 100% - 100% |
| S+20%T | 59% - 69% | 96% - 94% | 84% - 86% |
| S+20%T | 44% - 54% | 81% - 79% | 69% - 71% |
| S+20%T | 29% - 39% | 66% - 64% | 54% - 56% |
| S+20%T | 14% - 24% | 51% - 49% | 39% - 41% |

The range of value presented correspond to transmission rate range of 5% - 8%, thus fixing face mask and physical distancing at reported levels.
S: symptomatic testing, U: Mass test, T: trace and test, dT: delayed trace and test.

If vaccine coverage (*V*) is below 53% (the threshold noted above), it would be necessary to also reduce the population size (Table 2). If vaccine coverage (*V*) is between 38% and 53%, 23% and 38%, or 8% and 23%, in addition to mass tests at 33% per day, it would be necessary to maintain a population size threshold 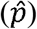 of at most 90%, 75%, or 60% on average, respectively, (Table 2) and the unvaccinated continue to use face masks and maintain physical distancing at current compliance levels. Note: the population size threshold noted here is the average of the values reported for contact rate (*C*) of 16 and 24 in Table 2.

Instead of adding mass tests only, addition of trace and tests only to symptom-based testing at the lowest rate of 10% (or highest rate of 50%) will also be sufficient to keep the population size on campus at 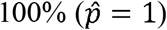 if at least 84% to 95% are vaccinated (or 42% to 63% are vaccinated) (Table 2). If vaccine coverage (*V*) is below 42% it would be necessary to also reduce the population size, keeping it to at most 89% on average if vaccine coverage is between 27% and 42%, and to at most 75% on average if vaccine coverage is between 12% and 27%. Considering that 50% trace and test, equivalent to 2 days from infection to isolation is a very tight timeline, which may be more feasible only with digital tracing, we also evaluated at a maximum of 20% trace and test, equivalent to 5 days from infection to isolation. This level of 20% trace and test only will be sufficient to keep the population size on campus at 100% if vaccine coverage is at least 74% to 84%. If vaccine coverage is below that, it will also require a reduction in population size, e.g., to 75% on average if only 44% to 54% of the population are vaccinated (Table 2). All the above scenarios for trace and tests also correspond to the continued use of face masks and physical distancing at least at current compliance levels (transmission rate of 5% to 8%). The combinations of testing and vaccination coverage under the full range of transmission rates are presented in Supplemental Appendix Table S5.

The total cases of infections and deaths over a 90-day semester if a fully unvaccinated population is on campus (contact rates of 16 to 24 per person per day as reported for regular face-to-face instructions [42,43]) suggest an exponential growth in infections in most testing scenarios, even if face mask and physical distancing are used at levels reported during the pandemic (transmission rates of 5% to 8%) (Supplemental Appendix Table S6). With contact rate of 6 to 8 per person per day (corresponding to reported numbers when several universities moved to partial or full remote instructions [35,36]) and use of facemask and physical distancing at levels reported during the pandemic, an exponential growth in infections was prevented with the following testing scenarios: 33% per day mass test only, at least 33% trace and test only, any of the combination tests, and any of the delayed combination tests (Table 3). In these suitable scenarios, the peak number of trace and tests, per 10,000 persons, varied from 2 to 64 per day, and the peak number of quarantines, per 10,000 persons, varied from 3 to 26 per day (Table 3).

**Table 3.** Suitable testing options for effective control of a disease outbreak keeping contact rates at reported levels ^† ‡^.

| Testing | Number infected (per 10,000 persons) |  | Peak trace and tests per day (per 10,000 persons) |  | Peak quarantine per day (per 10,000 persons) |  |
| --- | --- | --- | --- | --- | --- | --- |
| Transmission rate ($p$) □--> | 5% | 8% | 5% | 8% | 5% | 8% |
| S+33%U | 23 (20, 27) | 44 (32, 67) | 0 (0, 0) | 0 (0, 0) | 4 (3, 5) | 7 (5, 13) |
| S+33%T | 30 (25, 36) | 60 (42, 89) | 14 (9, 17) | 22 (14, 28) | 12 (9, 14) | 24 (18, 33) |
| S+50%T | 23 (21, 26) | 34 (29, 41) | 20 (12, 24) | 29 (19, 36) | 10 (9, 13) | 18 (14, 22) |
| S+5%U+50%T | 19 (17, 21) | 25 (22, 29) | 8 (7, 13) | 16 (10, 19) | 6 (5, 7) | 9 (8, 12) |
| S+10%U+50%T | 17 (16, 19) | 22 (20, 24) | 6 (5, 9) | 8 (7, 10) | 5 (4, 5) | 7 (6, 8) |
| S+20%U+50%T | 16 (15, 16) | 19 (18, 21) | 4 (3, 4) | 4 (4, 5) | 4 (3, 4) | 5 (4, 5) |
| S+25%U+50%T | 15 (15, 16) | 18 (17, 20) | 3 (2, 4) | 3 (3, 3) | 3 (3, 4) | 4 (4, 4) |
| S+33%U+50%T | 15 (14, 16) | 17 (16, 18) | 2 (2, 2) | 2 (2, 3) | 3 (3, 3) | 4 (3, 4) |
| S+5%U+50%dT | 30 (26, 34) | 45 (38, 55) | 26 (16, 32) | 46 (36, 64) | 15 (12, 18) | 26 (21, 34) |
| S+10%U+50%dT | 24 (22, 27) | 34 (29, 40) | 11 (9, 18) | 24 (15, 32) | 10 (8, 12) | 17 (14, 21) |
| S+20%U+50%dT | 19 (18, 21) | 25 (22, 28) | 4 (3, 8) | 10 (6, 10) | 6 (5, 7) | 9 (8, 11) |
| S+25%U+50%dT | 18 (17, 19) | 22 (20, 25) | 3 (3, 3) | 6 (4, 8) | 5 (5, 5) | 7 (6, 9) |
| S+33%U+50%dT | 17 (16, 17) | 19 (18, 22) | 2 (2, 2) | 3 (2, 6) | 4 (4, 4) | 6 (5, 7) |

S: symptomatic testing, U: Mass test, T: trace and test, dT: delayed trace and test
†Reported value of contact rate is on average between 6 and 8 per person per day under remote instructions [35,36]; Using our estimations, this would correspond to population size of about 31% and 42%. Results in the table correspond to this contact rate, presented as average (minimum, maximum)
‡ Reported contact rate is on average between 16 and 24 per person per day under face-to-face instructions [42,43]. None of the scenarios for this contact rate were suitable, and thus, are not presented in the table.
□ We defined a testing scenario as suitable if there were no exponential growth in infections when transmission rates were 5% and 8% (corresponding to reported use of face mask and physical distancing [11,30]).

## Discussions

This work estimates, under varying combinations of mass test, trace and test, and transmission rate, the contact rate thresholds that would help efficiently control an infectious disease outbreak on residential university campuses in the United States. The metric typically used in the COVID-19 literature for evaluating testing strategies is the reproduction number *R*_0_, which combines the contact rate and transmission rate. As interventions that influence transmission rates are different than those that influence contact rates, separating these parameters help systematically evaluate metrics to inform epidemic control protocols on university campuses. In this study, we extracted four main metrics. First, the contact rate threshold among non-essential workers after accounting for the higher contact rate among essential workers, which could help inform the size of social circles at the individual-level and schedule group activities such as in labs and offices. Second, population size threshold, i.e., the maximum proportion of the actual population size, which could help university-level activity decisions such as the fraction of classes that should be moved to remote instruction. Third, the threshold values for vaccination coverage for the campus to return to normal, i.e., the minimum vaccination coverage for having 100% of the population back on campus, which would help plan for the period post introduction of vaccines. Fourth, the threshold values for population size if vaccine coverage is below required thresholds, which could help decisions in the event that vaccines are not widely available that coverage (proportion vaccinated) is not at levels sufficient to fully resume normal activities. The fourth metric could especially be useful in the transitionary phase to normality (until vaccines become fully available) and where the results suggest lowering the population size by just a small number, which could be achieved by moving only a few classes online, such that, the overall population density on campus on any given day is lower but most students have most (if not all) of their classes as face-to-face.

While the implementation of the decisions related to the above metrics are driven at the university-level, adherence and feasibility to use of interventions such as face mask and physical distancing could vary by individual behaviors [37,39,40]. By separately modeling contact rates and transmission rates in this study, we extracted results corresponding to transmission rates (of 5% to 8%) that match reported behaviors for face mask use and physical distancing [11,30], and thus evaluated the university-level decisions under these adherence or feasibility ranges.

Our analyses suggest that implementing only testing, only face mask use and physical distancing, or only population size reductions will not be sufficient, but require combinations of these interventions to successfully control an outbreak on university campuses. Further, in the absence of vaccinations, at reported levels of face mask and physical distancing, testing alone without reducing population size would also not be sufficient to control an outbreak. This suggests that university campuses have high population densities that, for effective control of highly virulent infections such as SARS-CoV-2, it would require reducing the population size such as through remote learning.

Although individual interventions are not sufficient, there are multiple choices for combinations of interventions to choose from if vaccines are unavailable. If, along with continuing face mask and physical distancing at current levels, the population size is kept to at most 34% (or 44%) of the actual population size, mass tests only of 25% (or 33%) per day would help control an outbreak (Table 1). The choice between mass tests of 25% per day vs. 33% per day should consider the costs of a greater proportion remote learning (quantitative and qualitative costs) vs. costs of both testing more often and testing a larger population.

An alternative to mass tests only would be trace and test only, along with continuing face mask and physical distancing at current levels and reducing population size. Trace and test only would also be sufficient at rates of 33% (or 50%) if population size is kept to at most 39% (or 52%) (Table 1). These population size range are close to the 34% (or 44%) reported above for 25% (or 33%) per day mass tests only. Trace and test of 33% and 50% correspond to 3 days and 2 days, respectively, from the time an infected person makes contact with an individual to effective isolation of that individual. Feasibility of this short turnaround times would determine the choice between use of mass test vs. trace and test. Turnaround times are expected to be shorter with digital contact tracing, such as smart phone apps, compared to manual tracing, and feasibility and adoption of apps could be higher among university students than general population. However, studies related to its feasibility and adoption followed by adherence to isolation, among other issues such as privacy and alternative digital technologies are only recently emerging [44–47]. Our results also suggest that, if these turnaround times are not achievable and further if there are any delays in trace and test initiation, then trace and test alone is not favorable (none of the delayed trace and test were feasible (Table 2)) and should instead adopt either mass tests only or mass test with trace and test. Use of mass test with trace and test could improve trace and test due to potential early diagnosis of index persons. Our results suggest that, if mass tests can increase trace and test to 50% (within two days from contact to isolation), there is more flexibility in trade-offs between mass test rates and contact rate thresholds, and thus more flexibility in population size (Table 2).

In the event that vaccines are available, the full population can be back on campus and resume normal activities provided at least 95% of the population is vaccinated (Table 2). If vaccine coverages are lower than 95%, resuming normal activities with the full population size on campus would require additional asymptomatic testing, with the level of testing depending on vaccine coverage. Mass tests of at least 25% per day would be sufficient if vaccine coverage is at least 70%, or mass tests of at least 33% per day would be sufficient if vaccine coverage is at least 59%. If vaccine coverage is below 59%, to control an outbreak, in addition to mass tests at 33% per day, it would also require lowering the population size to 90%, 75%, and 60%, if vaccine coverage is at 46%, 31%, and 16%, respectively (Table 2).

Corresponding to the reported compliance to face mask and physical distancing and reported contact rates of 6 to 8 per person per day (a population size of 31% to 42% as per our estimations), from surveys [35,37,39] conducted over the year 2020 when universities transitioned a large proportion of classes to remote instructions and vaccines were unavailable, our results suggest the need for at least 33% mass test only or 33% trace and test only (Table 3). Scenarios that did not meet these criteria led to exponential growths in infections. These results generally match observed cases over the Fall 2020 semester, where several campuses saw cases into the thousands within the first two weeks of opening and were able to quickly control the spread within two to three weeks by temporarily transitioning to remote instructions [48]. While the universities were able to effectively control the outbreak quickly, it was also observed by this study [48] that the infections rapidly spread into the neighboring community, which were less successful in controlling the spread. Therefore, we believe, results obtained from our study, which set tight tolerance levels on infection cases, would be beneficial for developing epidemic response plans that consider the interests of the broader community. Our results also suggest that, with asymptomatic testing only, it would be necessary to have a vaccination coverage threshold of >95% for a university to fully return back to normal. This threshold is much higher than the typical 70% to 80% range used for herd-immunity in the literature for the general population [49], to a small extent because of setting a tighter tolerance but to a large extent because of the higher population density characteristic of university campuses. The latter can also be observed in *R*_0_ values estimated for universities, which in some instances went above 10 even with online instructions [48], while the herd-immunity in the general population is approximately calculated as 1 − 1/ *R*_0_ using a *R*_0_ of 3.5.

Our work is subject to limitations. Our model is deterministic. We used an average contact rate for all persons in order to estimate threshold values that could help inform university-level decisions. We did not model contact rates to be representative of actual expected networks between individuals. We did not explicitly model other interventions that could reduce transmission rate such as controlled ventilation, filtering air and controlling air flow, which are likely to impact transmissions [50]. The transmission rates also have a large range of uncertainty due to varying individual behaviors, the data used for streamlining the analyses in this study are based on limited data availabilities, however, the extrapolations over the wide range of transmission rates could be utilized. We did not model false positives for any of the testing scenarios and thus susceptible persons immediately return back to susceptible compartment after testing. We did not model other flu like illnesses and thus we did not assess the additional healthcare resource needs such as testing and quarantining because of similarity in symptoms with COVID-19. In estimation of vaccination thresholds, we did not consider the natural immunity developed among persons who may have been infected previously. The estimation of vaccination thresholds assume that the virus is still prevalent in the larger community and thus there is a chance of the infection entering the population, such as through local or global travel. We assume that the population density is similar across university campuses with contact rates between 16 and 24, and thus, this assumption should be considered when generalizing to campuses.

In conclusion, the results from this study could be used to collectively inform decisions related to testing, population size reductions through remote instructions, size of social circles, personnel scheduling in labs and offices, under scenarios of both unavailability or partial availability of vaccines, and within the observed levels of compliance to face mask use and physical distancing. The analyses conducted here specifically streamlined the results to the COVID-19 disease caused by the SARS-CoV-2 virus. However, given the wide range of transmission rates evaluated here, which were based on results from a meta-analysis study that evaluated SARS-CoV-2 and other viruses of similarly high virulence [11], broader observations from this study could be extrapolated for use in early stages of new outbreaks of similar viral respiratory infections with similar incubation periods, [24] where non-pharmaceutical intervention options such as face masks, physical distancing, remote instructions, and testing are the suitable options. As was the case at the time of conducting this study, in the early stages of an outbreak, there is uncertainty in the baseline transmission rate, efficacy of face mask use and physical distancing [11]. Thus, the results over the range of transmission rates might only serve as a preliminary guide, until more information becomes available for more streamlined analyses.

## Data Availability

All data generated or analyzed during this study are included in this published article.

## Financial Disclosure Statement

CG, XZ, and HT were partially funded by the National Science Foundation #1915481. The funders had no role in study design, data collection and analysis, decision to publish, or preparation of the manuscript.

## Acknowledgements

We would like to acknowledge Sonza Singh, Shifali Bansal, Seyedeh Nazanin Khatami, and Arman Mohseni Kabir for their assistance in data collection in initial stages of the study, and Dr. Laura Balzer, Dr. Michael Ash, and Dr. Hari Balasubramanian for their comments and inputs.

## Notes

### Competing Interest Statement

The authors have declared no competing interest.

### Funding Statement

Chaitra Gopalappa, Xinmeng Zhao, and Hanisha Tatapudi were partially funded by the National Science Foundation #1915481. The funders had no role in study design, data collection and analysis, decision to publish, or preparation of the manuscript.

