## Appendix for "Threshold analyses on combinations of testing, population size, and vaccine coverage for COVID-19 control in a university setting"

| **Parameter** | **Description**  (notations in main paper Methods) | **Age Range** | **Median** | **Range (LB)** | **Range (UB)** | **Source** |
| --- | --- | --- | --- | --- | --- | --- |
| Baseline transmission rate | $p$ |  | 14.3 | 11.6 | 17 | [1–4] |
| Latent period duration (days) | Not infectious; not symptomatic ($days_{L}$) |  | 2.52 |  |  | [5] |
| Infectious incubation period (days) | Infectious and not symptomatic ($days_{incub}-days_{L}$) |  | 3.5 |  |  | [6,7] |
| Incubation period duration (days) | $days_{incub}$ |  | 5.4 |  |  | [6,7] |
| Proportion of cases that never show symptoms | $prop_{asymp}$ |  | 0 |  |  | [7] |
| Time from onset of symptoms to recovery (days) | $days_{IR}$ |  | 7 |  |  | [7] |
| Base testing rate | $a_{B}$ (Rate for generating 65% symptom-based diagnosis) |  | 0.2 |  |  | [6–8] |
| Time from symptoms to hospitalization (days) | $days_{Q_{I}H}$ |  | 3.5 |  |  | [9] |
| Time from diagnosis to recovery (days) | $days_{Q_{I}R}$(same as $days_{Q_{I}H}$) |  | 3.5 |  |  | [9] |
| Proportion of cases that are hospitalized (also used as proxy for proportion of cases that are severe) | $prop_{hosp}$ (same for $prop_{severe}$) | 0–19 | 2% | 1.6–2.5 |  | [10] |
|  |  | 20–44 | 14% | 14.3–20.8 |  |  |
|  |  | 45–54 | 24% | 21.2–28.3 |  |  |
|  |  | 55–64 | 25% | 20.5–30.1 |  |  |
|  |  | 65–74 | 35% | 28.6–43.5 |  |  |
|  |  | 75–84 | 44% | 30.5–58.7 |  |  |
|  |  | ≥85 | 32% | 31.3–70.3 |  |  |
|  |  | Total | 12% | 20.7–31.4 |  |  |
| Duration of hospitalization for those who recover | $days_{HR}$ | 0-9 | 2 |  |  | [11] |
|  |  | 10-19 | 1.8 |  |  |  |
|  |  | 20-29 | 2.5 |  |  |  |
|  |  | 30-39 | 3.7 |  |  |  |
|  |  | 40-49 | 3.9 |  |  |  |
|  |  | 50-59 | 3.8 |  |  |  |
|  |  | 60-69 | 4.3 |  |  |  |
|  |  | 70-79 | 4.6 |  |  |  |
|  |  | 80-89 | 4.4 |  |  |  |
|  |  | 90-100 | 4.8 |  |  |  |
|  |  | Overall | 3.9 | 2.4 |  |  |
| Proportion of cases that recover | $prop_{recover}$ |  | Male;Female | Both |  | [11,12] |
|  |  | 0-9 | 100%;100% | 98% |  |  |
|  |  | 10-19 | 100%;100% | 97% |  |  |
|  |  | 20-29 | 93%;98% | 96% |  |  |
|  |  | 30-39 | 95%;98% | 93% |  |  |
|  |  | 40-49 | 92%; 97% | 89% |  |  |
|  |  | 50-59 | 88%; 93% | 82% |  |  |
|  |  | 60-69 | 81%; 88% | 70% |  |  |
|  |  | 70-79 | 64%; 73% | 51% |  |  |
|  |  | 80-89 | 39%; 52% | 12% |  |  |
|  |  | 90-100 | 36%; 54% | 12% |  |  |
| Duration of hospitalization for deaths | $days_{HD}$ | 0-9 | 2 |  |  | [11] |
|  |  | 10-19 | 1.8 |  |  |  |
|  |  | 20-29 | 4 |  |  |  |
|  |  | 30-39 | 2.8 |  |  |  |
|  |  | 40-49 | 5.6 |  |  |  |
|  |  | 50-59 | 5.9 |  |  |  |
|  |  | 60-69 | 5.7 |  |  |  |
|  |  | 70-79 | 5 |  |  |  |
|  |  | 80-89 | 3.9 |  |  |  |
|  |  | 90-100 | 3 |  |  |  |
|  |  | Overall | 4.8 | 2.3 |  |  |

**Table S1. Epidemic parameters used for the compartmental simulation model**

|  | **Relative risk [2]** | | | **Lower (baseline)** | | | **Median (baseline)** | | | **Upper (baseline)** | | |
| --- | --- | --- | --- | --- | --- | --- | --- | --- | --- | --- | --- | --- |
|  |  |  |  | **11.60%** | | | **14.00%** | | | **11.60%** | | |
| **Intervention** | Lower | Median | Upper | Lower | Median | Upper | Lower | Median | Upper | Lower | Median | Upper |
| **Facemask** | 0.40 | 0.56 | 0.79 | 4.6% | 6.5% | 9.2% | 5.6% | 7.8% | 11.1% | 6.8% | 9.5% | 13.4% |
| **3ft distancing** | 0.26 | 0.49 | 0.93 | 1.5% | 2.8% | 5.3% | 1.8% | 3.4% | 6.4% | 2.2% | 4.1% | 7.7% |
| **6ft distancing** | 0.10 | 0.20 | 0.41 | 1.2% | 2.3% | 4.8% | 1.4% | 2.8% | 5.7% | 1.7% | 3.4% | 7.0% |

**Table S2. Transmission rate estimates; Baseline values from [1–4], and relative risks from [2];**

| **Reference** | **Type of study** | **Timeline** | **Location/setting** | **Summary for number of contacts** | **Summary of face mask and social distancing compliance** |
| --- | --- | --- | --- | --- | --- |
| *University setting* | | | | | |
| [13] | Survey of students on the likelihood to engage in COVID-19 disease mitigation behaviors | During pandemic in early August, 2020 | United States/Kansas State University |  | 70.06% participants responded definitely will and 20.82% responded possibly will for usage of face mask in public spaces. |
|  |  |  |  |  | 78.2% participants responded definitely will and 16.3% responded probably will for use of face mask in classrooms. |
|  |  |  |  |  | 48.95% participants responded definitely will and 38.86% responded possibly will for observing 6 ft social distancing in public spaces. |
|  |  |  |  |  | 47.39% participants responded definitely will and 41.22% responded probably will for observing 6 ft social distancing in classrooms. |
| [14] | Survey of students regarding COVID related experiences | During early pandemic between April 25–30, 2020 | United States/University students across the country | Average number of contacts is 12.7. | Approximately 50.8% participants reported always wearing a face mask or covering in public. |
|  |  |  |  | Contacts ranged from 0 up to 100 |  |
|  |  |  |  | 4.2% were uncertain about number of contacts. |  |
| [15] | Modelling study to determine whether in-person instruction is safe to continue during the pandemic | During pandemic | United States/University setting | The model generates an average of 11 traceable and 8 nontraceable contacts per person per day when there are face-to-face classes and social distancing is not being exercised. | The transmission probabilities in the study were reduced by 50% in lieu of implementing non-pharmacological interventions (like mask wearing). |
| [16] | Survey conducted to determine the spread and frequency of protective behaviors, emotional and anxiety status. | During pandemic in early April, 2020 | Turkey/University students across the country |  | 50% reported wearing protective gloves and masks. |
| [17] | Survey conducted to understand social distancing and contact behavior. | Before pandemic between March and May 2003 | Belgium/University setting | The number of conversational contacts is 18.1 on weekdays and 12.3 on weekends. |  |
| [18] | Survey conducted to understand social distancing and contact behavior. | Before pandemic between 2003 and 2006 | Germany/University setting | The median number of conversational contacts varied between 6 and 11 for weekdays and weekends, respectively. |  |
| [19] | Survey conducted to understand social distancing and contact behavior. | Before pandemic between June and July 2004 | Germany/School setting | A mean number of contacts per day for children was 25.1. |  |
|  |  |  |  | The mean number of contacts per day for adults was 7.5. |  |
| [20] | Online survey of contacts, behavior, and symptoms | During pandemic between September and November, 2020 | United Kingdom/University of Bristol (UoB) | The median number of contacts on the day before the interview was 2. | Residential students were asked not to host non-residents in their household. |
|  |  |  |  | The interquartile range was between 1 and 5; the mode was 1, and the mean was 6.1. | Residential students were allowed to meet others outside of their household. |
|  |  |  |  | Among all participants, 8% had 20 contacts. | Students were asked to conform to government suggested social distancing guidelines and other mitigation measures (like face coverings) |
|  |  |  |  | Among students, 57% of student contacts were other UoB students/staff. |  |
|  |  |  |  | The mean number of contacts reported by students for the previous day was 6.1, with a median of 2. |  |
| [21] | Modeling study that uses data of online survey of contacts, behavior, and symptoms | During pandemic but contact data obtained from pre-pandemic survey conducted in September 2010 | United Kingdom/University of Bristol (UoB) | Students were assumed to living in groups with a maximum of 24 individuals. | The model reduced transmission probability by 25% for face covering and 50% for social distancing. |
|  |  |  |  | Intervention measures would reduce group size (i.e., number of students that share bathroom/kitchen facilities) from 24 to 20 or 14 students was seen as the least effective intervention. |  |
| [22] | Online questionnaire survey to determine knowledge, attitudes, and practices towards COVID-19 | During pandemic between May and June, 2020 | Japan/University students across the country |  | Approximately 52.1% of university students in Jordan and 98.0% of university students in China reported wearing a facemask when leaving home. |
|  |  |  |  |  | 86.9% of undergraduate students from Indonesia and 96.4% of Japanese students wore masks frequently in a crowded place. |
|  |  |  |  |  | The frequency of handwashing and mask-wearing were reported at a median of 96.4% |
| *Non-university setting* | | | | | |
| [23] | Survey for individual's perceptions about risk and behavior related to COVID-19 | During pandemic between May and June, 2020 | United Stated (six states in the U.S.: Colorado, Iowa, Louisiana, Massachusetts, Michigan, Washington)/Non-university setting |  | 66% participants reported always wearing a mask in public indoor spaces. |
|  |  |  |  |  | 46% participants reported always physically distancing outside home. |
| [24] | Multiple surveys conducted about COVID behavior | During pandemic between April and November 2020. | United States/Non-university setting |  | 77% participants reported to closely adhering to recommendations to wearing masks in November. |
| [25] | Survey to assess the impact of physical distancing policies on contacts | During pandemic between March and September, 2020 | United States/Non-university setting | A median of 2 contacts (0 non-household) in Wave 0, a median of 3 contacts (1 non-household) in Wave 1, a median of 3 contacts (1 non-household) in Wave 2, and a median of 4 contacts (2 non-household) in Wave 3. |  |
| [26] | Survey conducted to understand social distancing and contact behavior. | During early pandemic between March and April 2020 | United States/Non-university setting | Individuals who completely or mostly isolate have 5 contacts per day. |  |
|  |  |  |  | Individuals who don't attempt to isolate themselves have an average of 52 contacts. |  |
|  |  |  |  | Working adults have an average of 13.9 contacts and non-working adults have an average of 4 contacts. |  |
| [27] | Large-scale survey of social encounters | Pre-pandemic survey conducted in October 2010 | Great Britain/Non-university setting | According to the survey, individual-only contacts peaked at a maximum of 20. |  |
|  |  |  |  | The distribution of contacts was characterized by a lognormal body with a power-law tail and an exponent of 22.45 |  |
| [28] | Large-scale survey of social contacts | Pre-pandemic survey conducted in September 2010 | Great Britain/Non-university setting | The mean number of individual contacts is 7.97. |  |
|  |  |  |  | Mean total number of contacts is 26.75. |  |
|  |  |  |  | When adjusted for age and gender biases, mean individual contacts and total contact rise slightly to 8.28 and 28.50. |  |
| [29] | Population-based survey to assess mixing patterns in eight European countries | Pre-pandemic survey conducted between May 2005 and September 2006 | 8 European counties (Belgium, Germany, Finland, Great Britain, Italy, Luxembourg, Netherlands, and Poland)/Non-university setting | Mean contacts per person per day is 13.4. |  |
|  |  |  |  | German participants reported fewest daily number of contacts with a mean of 7.95 and Italians reported the highest number with a mean of 19.77. |  |
|  |  |  |  | 23%, 21%, 14%, 3%, and 16% of the reported contacts are made at home, at work, at school, while travelling, and during leisure activities, respectively when contacts were pooled together. |  |
| [30] | Survey conducted to evaluate the impact of government interventions on social contact patterns. | During pandemic between March and June, 2021 | Luxembourg/Non-university setting | Luxembourg, Italian, Belgian, British, and German residents reported and average of 17.5, 19.8, 11.8, 11.7, and 8 social contacts per day respectively, before the pandemic. | The average number of contacts per day as reported by participants was 7.1 contacts after the lockdown. |
|  |  |  |  | After the lockdown participants reported an average of 7.1 contacts per day on. | Participants reported that 61.7% of the total contacts occurred without a facemask, which is a mean of 4.9 contacts. |

**Table S3. Literature review of population behavior for number of contacts and compliance to face mask usage and social distancing for university and non-university setting (includes information from pre-pandemic and during pandemic surveys and model assumptions)**

**Table S4 (see Excel file): All combinations of testing, contact rates for non-essential workers, and population size for effective control of a disease outbreak in the absence of vaccines**

S: symptomatic testing, U: Mass test, T: trace and test, dT: delayed trace and test

‡ We assume 23% are essential workers and have a contact rate of 14 per day [14]

| **Shaded grey** | Infeasible scenarios: We assume a contact rate of 1 or below among non-essential workers (columns G to H) are infeasible, and thus corresponding rows (combinations of testing and population sizeare infeasible) |
| --- | --- |

† Contact rate threshold (per day): the average value for contacts per person per day to keep infections below the tolerance level. These reduced contact rates, from the original rates of 16 to 24, can be achieved through reduction in population size at the noted thresholds

**Table S5 (see Excel file). Combinations of testing, vaccine coverage, and population size for effective control of a disease outbreak**

S: symptomatic testing, U: mass test, T: trace and test, dT: delayed trace and test

**Table S6 (see Excel file). Epidemic outcomes under varying levels of testing and corresponding resource needs**

| S: symptomatic testing, U: mass test, T: trace and test, dT: delayed trace and test | |
| --- | --- |
| †Reported value of contact rate is on average between 6 and 8 per person per day under remote instructions [20,33]; Assuming contact rate is directly proportional to the population density, contact rate of 6 to 8 corresponds to reducing the population size to between 31% and 42% | |
| ‡ reported contact rate is on average between 16 and 24 per person per day under face-to-face instructions [17,27] | |
| ⁋ We defined a testing scenario as suitable if there were no exponential growth in infections when transmission rates were 5% and 8% (corresponding to reported use of face mask and physical distancing [1,2]) | |
| **Shaded grey** | Scenarios that are not suitable: We assume scenarios that lead to an exponential growth in infections are not suitable |
